# Neighborhood Social Vulnerability and Access to Expedited Partner Therapy Prescriptions: A Secret Shopper Audit Survey

**DOI:** 10.1101/2024.12.28.24319274

**Authors:** Rachel E. Solnick, Carmen C. Vargas-Torres, Alexis Guastello, Marissa Seldes, Kelsey Simpson, Patricia Mae Martinez, Michael Grant, Ethan Cowan

## Abstract

**Background:** Sexually transmitted infections (STIs) disproportionately impact populations with higher social vulnerability. Expedited Partner Therapy (EPT), which allows the treatment of partners without requiring a medical visit, reduces STI reinfection rates and expands treatment access for underserved groups. However, EPT remains underutilized, particularly in the electronic prescription era, which introduces logistical complexities. Previous studies highlight low pharmacist awareness of EPT, but few have assessed its real world availability in pharmacies or how accessibility varies by socioeconomic subcomponent of the Social Vulnerability Index (SVI).

**Objective:** This study aimed to evaluate pharmacist awareness and willingness to dispense electronic, nameless EPT prescriptions in New York City (NYC) pharmacies and examine how these outcomes vary by the socioeconomic factors of the pharmacy’s location.

**Study Design:** A cross-sectional study of 347 randomly sampled NYC pharmacies was conducted using a telephone-based secret shopper approach. Research associates posed as patients seeking to fill hypothetical EPT prescriptions to assess pharmacist awareness, willingness to dispense, and insurance acceptability. Multivariable logistic regression models evaluated the association between EPT awareness and willingness with SVI, adjusting for pharmacy type, neighborhood location, and local chlamydia rates.

**Results:** Among surveyed pharmacies, 40% (134/335) of pharmacists were aware of EPT, and only 30% (100/333) were willing to fill nameless prescriptions. Non-chain pharmacies were significantly less likely to be aware of EPT compared to national chains (34% vs. 54%, p=0.02). The most common dispensing approach was filling prescriptions under the index patient’s name (34%, 114/335), with most pharmacies (86%, 179/208) accepting insurance. Only 30% (100/333) of pharmacists were willing to dispense nameless EPT prescriptions. Most pharmacists’ methods of dispensing EPT prescriptions did not follow NY state EPT guidelines (54%, n=113). The most frequent reasons for refusal included unfamiliarity with EPT (62%, 66/107) and the incorrect belief that patient names were legally required (28%, 30/107). Adjusted regression showed increased odds of awareness of EPT in areas with the highest socioeconomic SVI quartile compared to the lowest quartile (odds ratio 3.7; 95% CI 1.4-10.8), though willingness to fill prescriptions did not differ by SVI (p=0.35).

**Conclusion:** Despite higher pharmacist awareness of EPT in more socioeconomically vulnerable areas, willingness to dispense nameless prescriptions remains low across NYC pharmacies. Independent pharmacies demonstrated particularly low awareness and engagement with EPT. These findings underscore the need for targeted pharmacist education, system-level interventions to streamline EPT dispensing, and enhanced training to ensure guideline adherence, particularly in high-need areas. Addressing these barriers could reduce STI disparities and improve public health outcomes.

## INTRODUCTION

Sexually transmitted infections (STIs) remain a persistent public health challenge in the United States(US). In 2023, over 2.4 million cases of chlamydia, gonorrhea, and syphilis were reported, with chlamydia accounting for more than 1.6 million cases.^1^ STIs continue to disproportionately affect marginalized populations, including racial and ethnic minorities, LGBTQ+ individuals, and socioeconomically disadvantaged groups. ^2,3^ Structural inequities, such as poverty, inadequate housing, and limited access to healthcare, contribute to these disparities.^4^

The health and economic consequences of STIs are substantial. Untreated infections can result in severe complications, including pelvic inflammatory disease and infertility, particularly in women.^5^ Additionally, STIs increase the risk of HIV transmission, further amplifying existing disparities in HIV prevention and outcomes.^6^ STIs also impose a significant financial burden, with the lifetime medical costs of chlamydia and gonorrhea exceeding $1 billion annually in the US.^7^ Addressing these issues requires effective strategies for both timely treatment and prevention.

Expedited Partner Therapy (EPT), endorsed by the Centers for Disease Control and Prevention, is an evidence-based strategy designed to facilitate treatment for the sexual partners of individuals diagnosed with STIs.^8^ By allowing healthcare providers to prescribe or dispense medications for partners without prior clinical evaluation, EPT helps reduce barriers to partner treatment and decreases rates of reinfection.^9,10^ However, despite being legal or permissible across the US, the implementation of EPT remains inconsistent, often due to limited pharmacist awareness and misconceptions about its regulations.^11,12^ Moreover, while electronic health records (EHRs) offer opportunities to streamline EPT prescribing, little is known about whether electronically prescribed EPT medications are fillable by pharmacists.

This study aims to evaluate the accessibility of electronically prescribed nameless EPT in New York City (NYC) pharmacies using a secret shopper methodology. Additionally, we examine whether disparities in EPT acceptance are associated with neighborhood-level socioeconomic social vulnerability, as measured by the socioeconomic subcomponent of the Social Vulnerability Index (SVI).^13^ Research shows communities with higher SVI scores experience worse health outcomes,^14^ greater barriers to healthcare access,^15^ and a higher burden of STIs.^16–18^ This association suggests that structural inequities, as demonstrated by higher SVI, may also hinder EPT implementation. We hypothesize that EPT accessibility will be limited across New York City pharmacies and that barriers will be more pronounced in areas with greater social vulnerability. Understanding these dynamics can help inform targeted interventions to improve EPT accessibility and reduce disparities in STI care.

## MATERIALS AND METHODS

### Study Design

We conducted a cross-sectional, observational audit survey utilizing a “secret shopper” methodology to evaluate the accessibility of electronically prescribed, nameless expedited partner therapy (EPT) prescriptions at retail pharmacies in NYC. The study aimed to quantify the proportion of pharmacies capable of dispensing EPT prescriptions and assess whether accessibility varied by neighborhood-level socioeconomic social vulnerability, as measured by the Centers for Disease Control and Prevention’s (CDC) Social Vulnerability Index (SVI).

### Procedures

A “secret shopper” study design in health services research involves a research assistant posing as a patient inquiring about healthcare resources, which promotes unbiased interaction with healthcare staff.^19^ Research assistants trained as pseudopatients simulated the role of the sex partners of STI-positive patients. A standardized script (**See Figure 1 and Script in Appendix A)** was developed and then reviewed by a consultant pharmacist for feasibility. The script was iteratively revised based on piloting with ten pharmacies. Using a standardized script, pseudopatients contacted pharmacies by telephone, stating that their partner was diagnosed with chlamydia and inquiring if an EPT prescription could be hypothetically sent electronically under the profile “Expedited Partner.” The prescription would include no personally identifiable information in compliance with New York State law. No prescription was actually sent in the scenario. To ensure feasibility, a pharmacist was consulted to review the script. The script was revised iteratively based on piloting from initial calls with ten pharmacies before data collection. Pseudopatients provided clarification and generic demographic details (e.g., the profile’s birthdate) upon request. Data collected during each interaction included whether the prescription was dispensed, the payment method (insurance versus out-of-pocket), provision of counseling (e.g., medication instructions, potential side effects, or sexual health recommendations), and total interaction time. If the pharmacist declined to fill the prescription, pseudopatients documented reasons for refusal, including pharmacist familiarity with EPT, technical barriers, or institutional policies. Data was recorded using a standardized Google form. Following the interactions, all pharmacies received debriefing letters detailing the study’s purpose to ensure ethical transparency.

**Figure 1.**
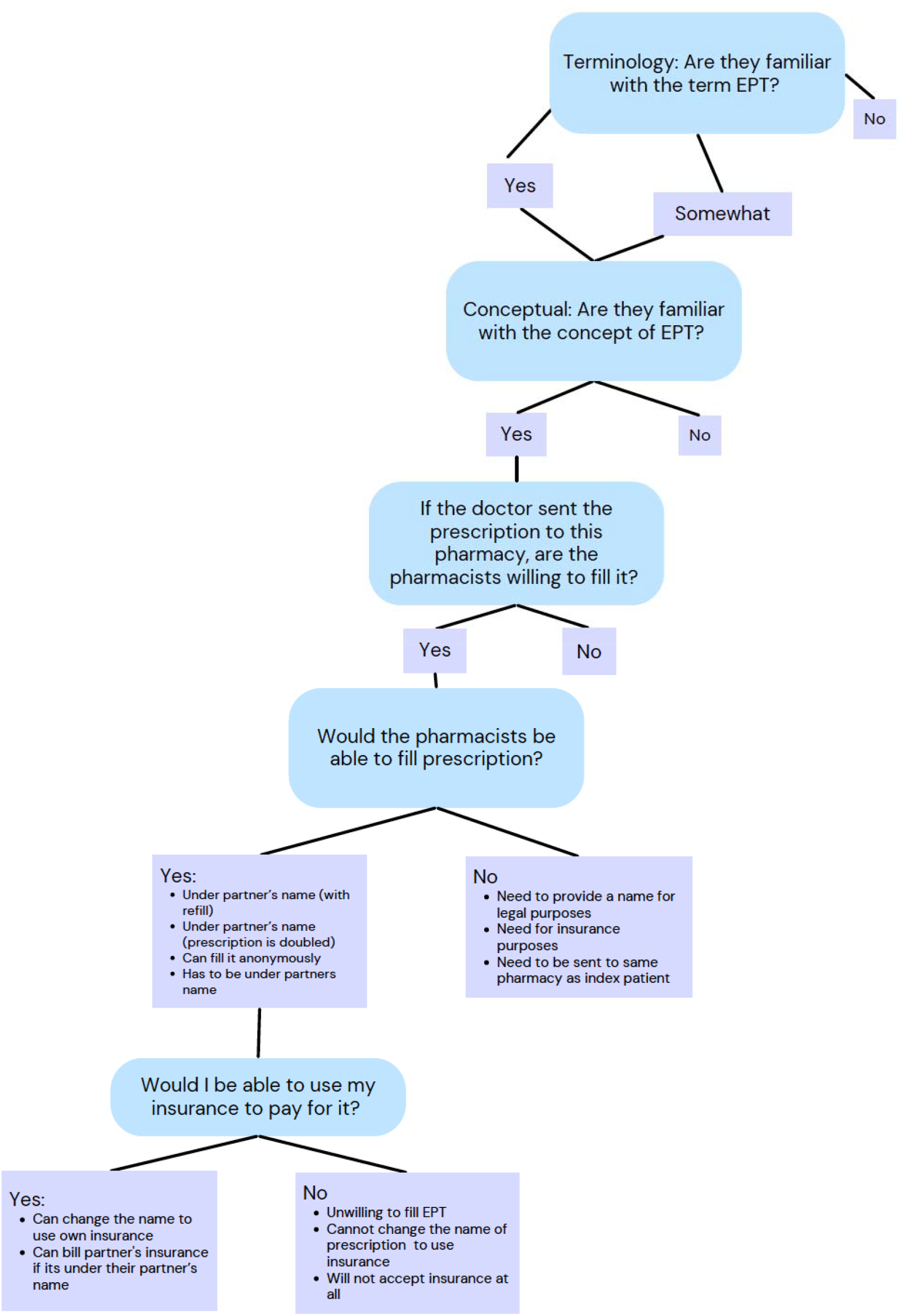
Script Flowchart for Pseudopatients to ask Pharmacists about EPT.

### Training of Pseudopatients

Pseudopatients underwent comprehensive training to ensure fidelity to the protocol. This included reviewing the standardized script and role-playing exercises. The principal investigator provided feedback to enhance consistency and ensure accurate data collection. To minimize bias, pseudopatients were blinded to the study’s secondary research objectives.

To begin the call, the pseudopatient asked the pharmacists about their familiarity with EPT prescriptions. The pseudopatient briefly explained the term if the pharmacist was unfamiliar with EPT. Willingness to dispense EPT as a nameless prescription (i.e., under the name “Expedited Partner”) was recorded. If the pharmacist was unwilling to fill the EPT prescription, the reason was documented.

Practice calls revealed that some pharmacists were willing to fill EPT prescriptions using methods that did not adhere to legal guidelines (i.e., filling it under the index patient’s name or insurance without the use of the term EPT in the prescription). These alternative methods were documented, and the pharmacy was considered aware of the term EPT but not the concept. Pharmacists were categorized as understanding the concept of EPT if they demonstrated a willingness to dispense EPT prescriptions in accordance with New York law and regulations– including allowing for either named or nameless prescriptions, for prescriptions to have “EPT” in the comments.^20–23^

For pharmacists willing to fill EPT prescriptions, the pseudopatient inquired about insurance use. Responses regarding insurance acceptance or estimated out-of-pocket costs were recorded. The call concluded with the pseudopatient stating they would discuss the prescription with their partner before asking the doctor to send a prescription to the pharmacy.

### Neighborhood-Level Social Vulnerability

Neighborhood characteristics were analyzed using the CDC SVI, which assigns composite scores to census tracts based on socioeconomic and demographic factors. Each census tract receives scores for individual variables within these themes and a composite score reflecting overall vulnerability. For this study, the socioeconomic subcomponent of the SVI was used, which includes variables such as the percentage of persons living below 150% of the federal poverty line, unemployment rate, percentage of housing cost-burdened occupied units (where at least 30% of income is spent on housing), percentage of persons aged 25 or older without a high school diploma, and the percentage of individuals without health insurance. Data for these measures were obtained from the CDC SVI Data and Documentation portal,^13^ with higher composite scores indicating greater levels of vulnerability. Pharmacies were linked to SVI data by census tract.

Additional covariates included in the logistic regression included pharmacy chain type (categorized as national, regional, or non-chain), neighborhood chlamydia incidence rates (reported as cases per 100,000 population and stratified into tertiles based on NYC Department of Health data). We adjusted for geographic variation by including neighborhood codes as categorical variables. Each neighborhood was represented by a unique code (n = 35) consisting of adjoining zip code areas, designated to approximate New York City Community Planning Districts.

### Pharmacy Sampling and Classification

Pharmacies within the catchment area of the Mount Sinai Health System, including Manhattan, Bronx, Brooklyn, and Queens, were randomly selected for this study. The sampling frame was developed using the New York State Board of Pharmacy’s comprehensive registry and supplemented with an online pharmacy locator database^24^ updated with data from Centers for Medicare and Medicaid Services (CMS) and containing approximately 3,000 pharmacies across four New York counties. From this larger sample, 300 pharmacies were randomly selected. To achieve a more balanced sample, additional pharmacies from the lowest SVI quartile were randomly selected and included to adjust for the skewed distribution of SVI values until we reached the final sample of 335.

Pharmacies were categorized as national chains (operating in >50% of U.S. states), local chains (limited geographic presence), or independents (not chain-affiliated). This classification allowed for stratification by pharmacy type, as prior studies^25^ have suggested that pharmacy characteristics may influence awareness and adherence to EPT policies.

### Study Outcomes

The primary outcomes were the hypothetical willingness to dispense EPT prescriptions and pharmacist awareness of EPT. Secondary outcomes were whether the primary outcomes varied by socioeconomic SVI quartile. Additional data included the pharmacist’s stated barriers to dispensing, approach to filling EPT (i.e., under the name of “expedited partner,” index patient, or the partner’s name), and whether insurance could be applied.

### Data Analysis

Descriptive statistics, including frequencies and proportions, were used to summarize primary and secondary outcomes. Bivariate analyses using chi-square tests assessed differences in EPT accessibility across pharmacy types, SVI quartiles, and neighborhood STI rates. Logistic regression models examined the association between EPT accessibility and SVI scores, adjusting for chlamydia incidence, pharmacy type, and geographic location. Mixed-effects models accounted for clustering by pharmacy type to produce robust estimates. Secondary analyses evaluated interactions between pharmacy type and specific SVI components, such as poverty and unemployment. Analyses were conducted with STATA Statistical Software (StataCorp version 16, College Station, TX). The Institutional Review Board at Mount Sinai Hospital reviewed and approved this project.

## RESULTS

Among the 335 pharmacies surveyed, 40% (n=134) of pharmacists reported awareness of EPT. **(TABLE 1)** Awareness was significantly associated with the willingness to dispense nameless EPT prescriptions. Pharmacists aware of EPT were more likely to dispense nameless prescriptions than those unaware (48% vs. 18%; p<0.001). However, neighborhood socioeconomic vulnerability was not associated with willingness to dispense nameless EPT prescriptions (p=0.35).

**TABLE 1.**
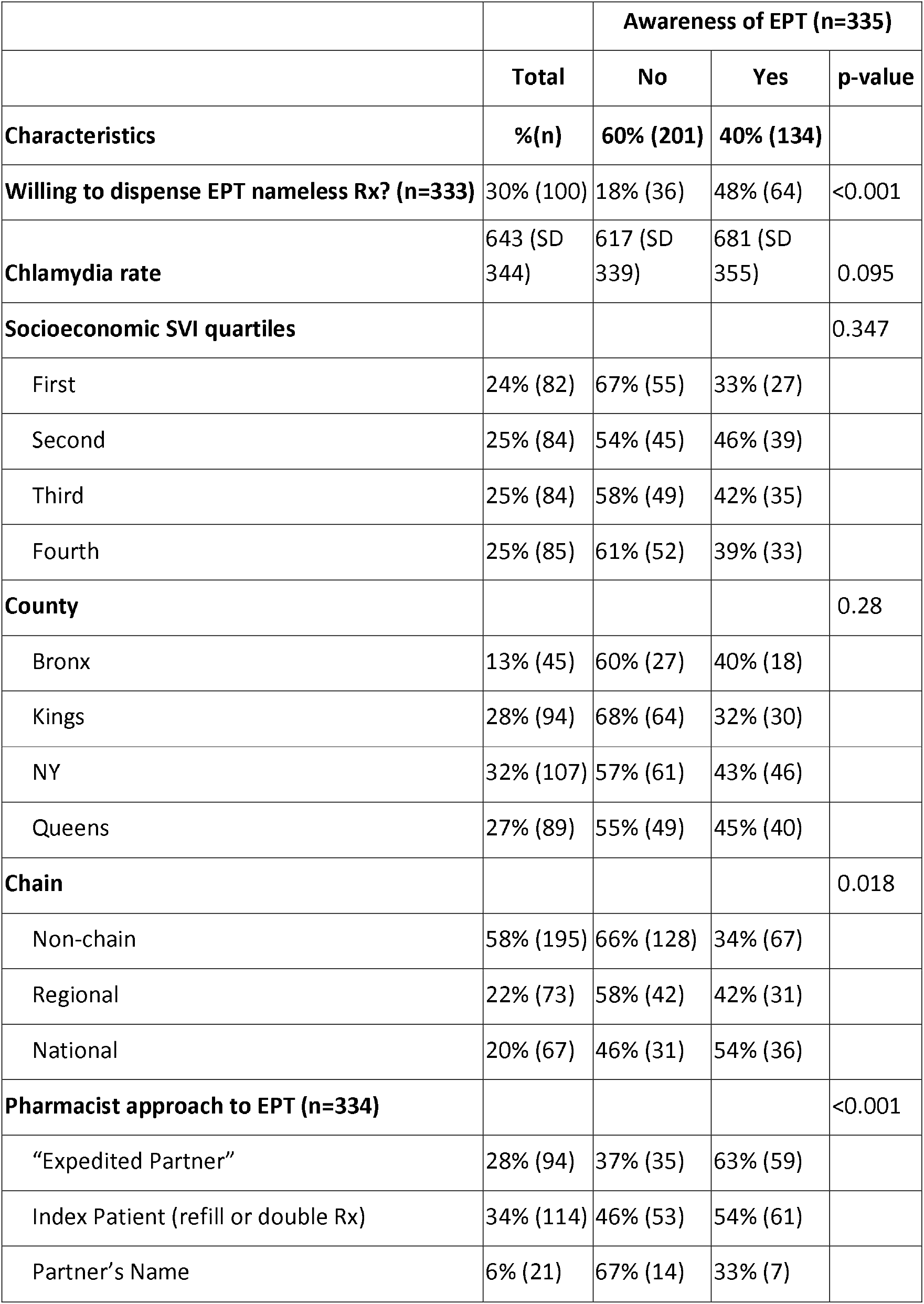
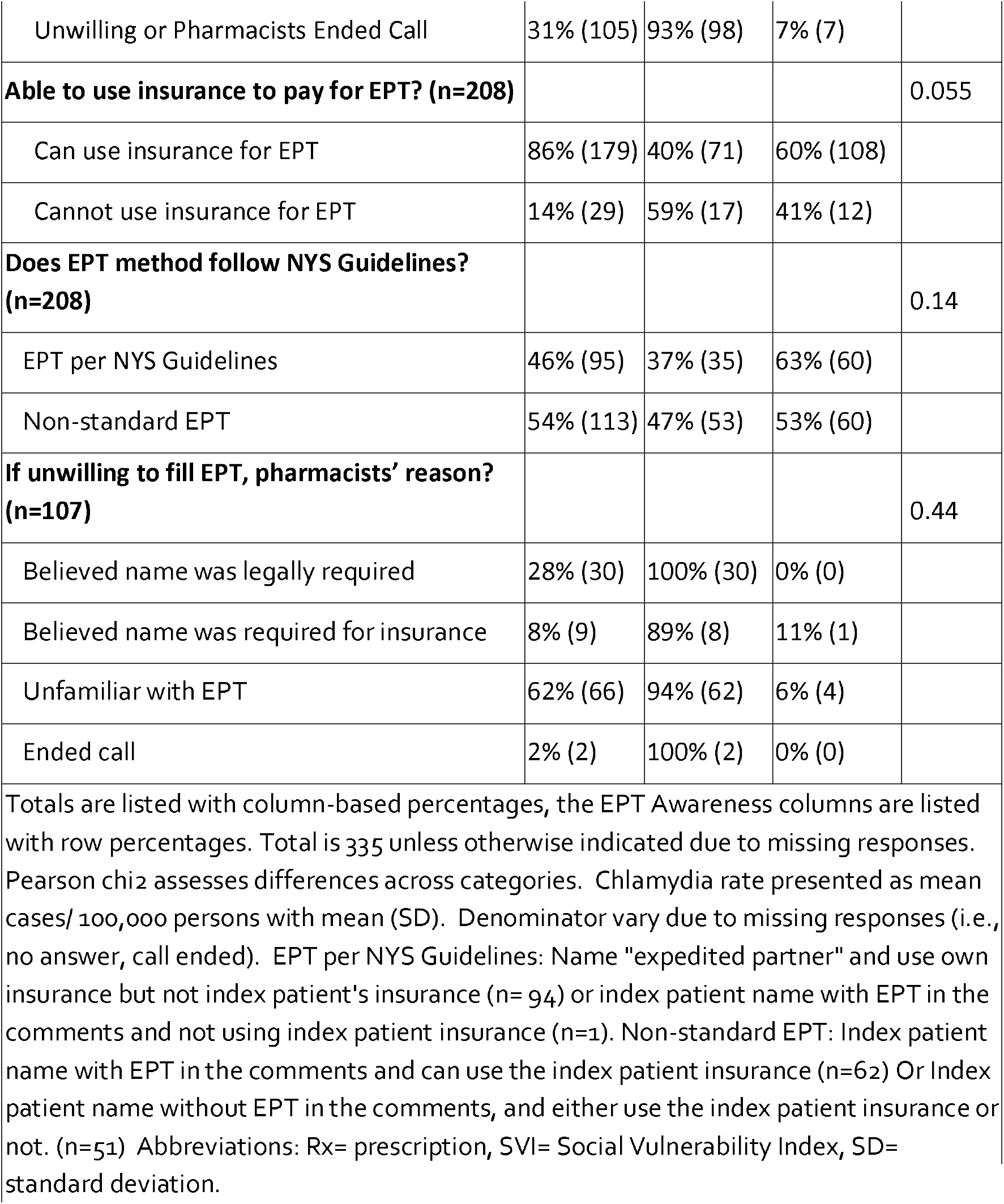
Pharmacist Awareness and Practices Regarding Expedited Partner Therapy (EPT) by Sociodemographic and Pharmacy Characteristics.

Pharmacist approaches to EPT prescriptions varied. Among pharmacists willing to dispense EPT, 28% (n=94) used the term “Expedited Partner,” 34% (n=114) used double prescriptions or refills under the index patient’s name, and 6% (n=21) required the partner’s name. In contrast, 31% (n=105) of pharmacists were unwilling to fill the prescription or terminated the call. Among those unwilling to fill the prescription, the most common reasons cited were unfamiliarity with EPT (62%, n=66) and the belief that a patient name was legally required (28%, n=30). Of the EPT prescriptions that the pharmacists would fill, less than half of pharmacists, 46% (n=95), adhered to New York State (NYS) guidelines for EPT prescribing.^21^ Pharmacists who were aware of EPT were more frequently NYS guideline-concordant (63%) compared to unaware pharmacists (37%)(p=0.14).

Insurance was usable in 86% (n=179) of potentially fillable EPT prescriptions and more frequently when the pharmacist was aware of EPT (60% vs 40%) (p=0.06). Neighborhood chlamydia rates were also not significantly associated with pharmacist awareness of EPT or willingness to dispense nameless prescriptions. The mean chlamydia incidence rate was slightly higher in areas where pharmacists were aware of EPT compared to those unaware (681 vs. 617 per 100,000; p=0.1). Borough-level analysis showed no significant differences in EPT awareness or dispensing across regions (p=0.28).

Logistic regression identified significant associations between pharmacist awareness of EPT and pharmacy type as well as level of neighborhood socioeconomic vulnerability. **(TABLE 2)** National chain pharmacies were more likely to report EPT awareness compared to non-chain pharmacies: adjusted odds ratio (aOR) 2.58, (95% confidence interval [CI] 1.31–5.06). Socioeconomic vulnerability, measured by SVI economic quartile, was positively associated with EPT awareness. Pharmacies with higher levels of socioeconomic vulnerability in the second, third, and fourth SVI quartiles were significantly more likely to report EPT awareness compared to those in the lowest quartile aOR 2.62, (95% CI 1.16–5.91); aOR 3.53, (95% CI 1.33–9.40); aOR 3.74, (95% CI 1.29–10.8), respectively. Chlamydia incidence rate tertiles were not significantly associated with EPT awareness. United Hospital Fund (UHF) codes served to adjust for regional variability and did not substantially affect the associations observed for pharmacy type or SVI. The logistic regression analysis for the outcome of pharmacists’ willingness to fill EPT prescriptions showed no significant associations between pharmacy type, chlamydia rate, or SVI quartile. (**TABLE 3)**.

**TABLE 2.** Factors Associated with Awareness of Expedited Partner Therapy (EPT) Prescriptions.

|  | Odds Ratio (Awareness of EPT) | [95% Conf. Interval] |  |
| --- | --- | --- | --- |
| **Pharmacy type** |  |  |  |
| Regional chain | 1.38 | 0.70 | 2.72 |
| National Chain | 2.58 | 1.31 | 5.06 |
| **Chlamydia rate (tertile)** |  |  |  |
| 2 | 3.27 | 0.45 | 23.74 |
| 3 | 2.94 | 0.37 | 23.09 |
| **SVI Economic quartile** |  |  |  |
| 2 | 2.62 | 1.16 | 5.91 |
| 3 | 3.53 | 1.33 | 9.40 |
| 4 | 3.74 | 1.29 | 10.83 |

Note: Pharmacy type: non-chain is reference group, SVI= Social Vulnerability Index, not displayed is United Hospital Fund (UHF) codes ORs which were also included in this model for space based adjustment

**TABLE 3.** Factors Associated with Willingness to Fill Expedited Partner Therapy (EPT) Prescriptions.

|  | Odds Ratio (Willingness to Fill EPT) | [95% Conf. Interval] |  |
| --- | --- | --- | --- |
| **Pharmacy type** |  |  |  |
| Regional chain | 0.80 | 0.43 | 1.48 |
| National Chain | 0.82 | 0.44 | 1.54 |
| **Chlamydia rate (tertile)** |  |  |  |
| 2 | 1.14 | 0.66 | 1.98 |
| 3 | 0.55 | 0.30 | 1.01 |
| **SVI Economic quartile** |  |  |  |
| 2 | 1.14 | 0.58 | 2.21 |
| 3 | 1.30 | 0.66 | 2.56 |
| 4 | 1.24 | 0.63 | 2.43 |

Note: Pharmacy type: non-chain is reference group, SVI= Social Vulnerability Index

## COMMENT

### Principal Findings

This study found that only 40% of pharmacists were aware of expedited partner therapy (EPT), and willingness to dispense nameless prescriptions remained low at 30%. EPT awareness was higher among national chain pharmacies compared to regional and independent pharmacies. However, adherence to New York State EPT guidelines was suboptimal, with only 46% of dispensed prescriptions meeting these standards. Notably, neighborhood chlamydia incidence did not significantly impact pharmacist awareness or willingness to dispense EPT.

We initially hypothesized that higher socioeconomic vulnerability would be associated with lower pharmacist awareness and willingness to fill EPT, potentially explaining a causal link between limited neighborhood resources and worse sexual health outcomes.^16^ Contrary to our expectations, higher socioeconomic vulnerability was associated with greater pharmacist awareness of EPT. This finding may reflect increased EPT utilization by patients in areas of greater need for alternative forms of healthcare which could heighten pharmacist familiarity with EPT. However, no association was observed between socioeconomic vulnerability and pharmacists’ willingness to fill EPT. This may suggest a lack of pharmacy-level resources— such as standardized workflow policies, electronic record interfaces, and time allocated for patient care—in these areas, which could limit pharmacists’ ability to dispense EPT.

### Results in the Context of What is Known

We also observed low adherence to state-level guidelines for processing EPT prescriptions, likely stemming from a lack of pharmacist familiarity with laws allowing the legal dispensing of nameless prescriptions. This finding aligns with a multistate study showing that vouchers for nameless EPT prescriptions were less likely to be successfully filled compared to those with patient names.^26^ Furthermore, when EPT is dispensed, it is often filled under the index patient, likely due to the logistical challenges of processing nameless prescriptions. This pattern was observed in a study of NYC pharmacies, where EPT was commonly prescribed using a double-dose approach.^27^ Similar gaps in regulatory knowledge have been reported elsewhere, such as in a Minnesota study, which identified legal and administrative concerns as significant barriers to EPT adoption among providers.^28^

This study highlights barriers to EPT implementation in NYC and contributes to a broader understanding of pharmacy-level challenges in urban settings. Consistent with prior studies, we found that fewer than half of NYC pharmacists knew EPT’s legality, mirroring findings from a 2014 NYC survey (42% awareness)^29^ and a 2022 Northeastern US survey (50% awareness).^30^ Limited pharmacist exposure to EPT contributes to a lack of familiarity with its procedures, particularly in lower-use areas, which may make it harder for patients to successfully fill prescriptions and further limit EPT use. Independent pharmacies in our study were less likely to report EPT awareness than national chains, potentially due to disparities in access to standardized training and institutional resources. Similarly, Qin et al. ^31^ identified pharmacy deserts in high-burden Baltimore neighborhoods, where limited geographic access compounded EPT implementation barriers. These findings suggest that while urban pharmacies in underserved areas may be more receptive to EPT, systemic challenges such as low awareness and cost variability persist.

Our findings of pharmacy-level barriers help further explain the persistent low implementation of EPT. For example, Stenger et al.^26^ found that on average, only 5.4% of patients with gonorrhea received EPT, with stark regional variation, ranging from 35.5% in Washington State and 0.04% in New York City. This potentially highlights the impact of state level policies and systemic interventions to change prescribing and filling practices. Similarly, a national study of EPT in emergency departments demonstrated significant gaps in knowledge and implementation, with only 38% of medical directors understanding how to prescribe it.^32^

### Clinical Implications

These findings reveal critical barriers to EPT implementation that could limit its public health impact. Low willingness to dispense nameless prescriptions restricts access to EPT, particularly in underserved areas which could most benefit. Given their higher awareness levels, national chain pharmacies may serve as focal points for implementing EPT interventions, but greater efforts are needed to standardize practices across all pharmacy types. Policymakers and professional organizations should focus on addressing legal and logistical barriers, such as pharmacist misconceptions about prescription requirements, insurance policies that do not cover partner prescriptions under EPT, and streamlining the process for electronic prescribing.

### Research Implications

Research is needed to evaluate the impact of electronic prescribing on either improving access or introducing new barriers to filling EPT. Future studies should explore strategies to improve pharmacist education on EPT, particularly in non-chain and regional chain pharmacies. Additionally, studies examining the cost-effectiveness of policy change that would allow index patients’ insurance to cover EPT–currently only legislated in California^33^– including its impact on reducing STI rates would provide critical evidence for policymakers and healthcare organizations looking to reduce health disparities in STIs.

### Strengths and Limitations

The strengths of this study include its use of a secret shopper methodology, which provided a realistic assessment of pharmacist practices, and the inclusion of socioeconomic vulnerability data, which offered insights into structural disparities. However, limitations include the hypothetical nature of the EPT prescription, which occurred via telephone call, which may have resulted in lower pharmacists’ willingness to fill the prescription, the potential variability in secret shopper interactions, which could introduce bias, and the study’s focus on a single metropolitan area, which limits generalizability. Additionally, this study did not directly examine pharmacists’ decision-making processes, which could provide further context for the observed gaps in EPT awareness and dispensing practices.

## CONCLUSIONS

This study highlights significant barriers to EPT implementation at the pharmacy level, including limited awareness, low willingness to dispense nameless prescriptions, and poor adherence to guidelines. These findings underscore the need for targeted educational initiatives, policy interventions, and the integrating of electronic prescribing systems to enhance EPT access and public health outcomes. Further research is needed to address geographic and structural disparities and optimize EPT implementation in high-need populations.

## Data Availability

All data produced in the present study are available upon reasonable request to the authors

## ACKNOWLEDGEMENTS

The authors gratefully acknowledge the Emergency Medicine Foundation Health Disparities Research Grant for their financial support of this study. Dr. Solnick was supported by grant K23MH136923-01 from the National Institutes of Health during this work. The authors also thank the Mount Sinai iCORE program for their development of a summer research program to help support the student research assistants. The EMF, iCORE, and NIH had no role in the design and conduct of the study; collection, management, analysis, and interpretation of the data; preparation, review, or approval of the manuscript; and decision to submit the manuscript for publication. We are grateful for the pharmacists who took the time to speak with us during the secret shopper calls. The authors declare no conflicts of interest related to this work.

## Disclaimer

The views expressed in this article are those of the authors and do not reflect the official position of the National Institutes of Health

## APPENDIX A. SCRIPT

The secret shopper interaction will be as follows:

### SCRIPT

#### START TIMING THE CALL

1. “Can I speak to a pharmacist please?”
  a. Potential response: “who’s calling?/why?”
    i. **How to respond:** “Im a patient, I just have a private question for the pharmacist?”
2. “Hey, my boyfriend recently got diagnosed with chlamydia and his doctor said I can get an EPT, or expedited partner therapy prescription that can be filled electronically, are you familiar with it?”
  a. Potential response: Pharmacists hedges ….”Sorta, A little bit”)
    i. **How to respond:** Then follow-up with a clarification question to make sure they know what EPT is “Its a way I can get an antibiotic for chlamydia without seeing a doctor, have you heard of that? “
  b. Potential response: have you been here before?
    i. No but I’m just calling to see if you will accept this prescription
3. IF THEY SAY NO/ASK A FOLLOW UP QUESTION: “so my boyfriend’s doctor said it was called expedited partner therapy and that the prescription for STI medication could be sent electronically for me even if I didn’t see the doctor?
  a. Potential Response: “Okay can I have your name / boyfriends name to look you up in our system?”
    i. **How to respond:** The doctor told my boyfriend that I can have this prescription filled anonymously, so it won’t be sent with either of our names. They told me that the name would be called “Expedited partner” and the birthday would be 1901. Is that something you can fill for me?
  b. Potential Response: “Who is the prescription for?”
    i. **How to respond:** “It’s for me, can you fill it for me?
  c. Potential Response: “what kind of medication is it?”
    i. **How to respond:** My boyfriend said its called doxycycline, will you be able to fill this type of prescription?
  d. Potential Response: “The doctor would not even send it that way” (they need to actually see the prescription to believe it could happen)
    i. **How to respond:** How would they send it?
  e. Potential Response: They would double the dose and state it’s for the partner
    i. **How to respond:** Should it have EPT anywhere on the prescription
4. IF THEY SAY YES: “Okay great, if his doctor sent the EPT prescription to this pharmacy, would it be possible to fill it?”
  a. Potential response: “Yes I can fill this for you.”
  b. Potential Response: “Yes but I need a name/date of birth to fill the prescription”

i. **How to respond:** “My boyfriend said his doctor said that I could just say it was a prescription that said EPT somewhere on it and that I wouldn’t need to provide a name/date of birth”
  c. Potential response: “No, I’m too busy”
  d. Potential response: “No, we can’t receive”
  e. Potential response: “No, we need the name for record keeping
  f. Potential response: “No, we need it for legal purposes
  g. Potential response: “No, we need it for insurance purposes
  h. Potential response “I have to check with my manager”
    i. **How to respond:** “Okay thank you for your time”
5. AFTER THEY AGREE TO FILL THE PRESCRIPTION: I just want to confirm that you will fill this even though It’s not going to have either my boyfriend or my name on it, It’s going to have “expedited partner” for the patient name? Would you still accept that?”
  a. Potential response “I have to check with my manager (includes, they ask the caller to call back”
  b. If yes, continue
6. IF THEY ARE NOT ABLE TO FILL IT UNDER EXPEDITED PARTNER: If you cannot fill the EPt prescription for me under the name expedited partner, how could you fill it for me?
7. Would I be able to use my own insurance to pay for the prescription?
  a. IF THEY SAY NO: Okay, can I pay for it out of pocket?
    i. IF YES: What will the cost be? …. Okay great I’m going to talk to my boyfriend first, thank you for your time.
  b. IF THEY SAY YES:
    i. Potential Response: the pharmacy will need your name on the script to bill the insurance
      1. **How to respond:** “Could you adjust the prescription to show that it is for me if I give you all the insurance info”
        a. IF THEY SAY NO, the system won’t allow us to do that.
        b. **How to respond**: “Is there any way to add my name to the prescription the doctor says so you can bill my insurance?”
        c. IF THEY SAY NO, still can’t do it
          i. **How to respond:** “Okay, can I pay the over the counter price then?”
          ii. ***AFTER THEY RESPOND END TIMING THE CALL***
    ii. Potential Response: the pharmacy can bill your boyfriend
      a. **How to respond:** “Just to clarify, I mean my own insurance, is that possible.”
  c. IF THEY SAY YES: Okay great I’m going to talk to my boyfriend first, thank you for your time.

